# Longitudinal Stability and Contextual Sensitivity of At-Home Salivary Dim Light Melatonin Onset (DLMO): A Four-Year N-of-1 Study

**DOI:** 10.64898/2026.08.03.26359048

**Authors:** Chris S. Schwartz, Steve W. Granger, Brittany M. Aldrich, Ellen R. Stothard, Robert J. Thomas

**Affiliations:** Salimetrics Research and Technology Center, Carlsbad, CA, USA; Rebis Health, Longmont, CO, USA; Department of Medicine, Beth Israel Deaconess Medical Center, Boston MA

## Abstract

At-home collected salivary Dim Light Melatonin (DLMO) assessments have demonstrated agreement with laboratory-determined DLMO estimates, but key translational gaps remain. We compiled 32 at-home salivary DLMO assessments obtained over four years from a healthy adult male (35– 40 years old) with stable sleep-wake patterns to characterize the magnitude of behavioral, environmental, and pharmacologic influences on DLMO and melatonin profile morphology at the individual level. The series included 10 assessments conducted under standardized dim light conditions to establish a baseline pattern, 21 assessments following controlled contextual manipulations (including exogenous melatonin, ambient and bright-light exposures, blue-blocking glasses, diazepam, behaviorally delayed bedtime scheduling, melatonin-rich foods, magnesium, caffeine, alcohol, late-evening anaerobic exercise), and one seasonal photoperiod comparison. Across a series of 10 baseline assessments, DLMO timing demonstrated high intra-individual stability (range: 7:13 PM–8:43 PM; SD = 28 min), and peak melatonin levels were highly consistent (M = 13.26 pg/mL, SE = 0.46 pg/mL). Within this individual, 5-day magnesium supplementation (3.6 mg/kg per day), caffeine (300 mg), alcohol (6oz of 80-proof), melatonin-rich foods, or intense anaerobic exercise during the DLMO assessment window produced negligible deviation in DLMO timing or changes in peak concentrations. In contrast, continuous bright light (∼1500 lux) during the established pre-onset interval suppressed melatonin production and obscured central circadian phase estimation, while bright light exposure after melatonin onset also produced steep declines in melatonin levels. Diazepam (0.12 mg/kg per day, over a 5-day period) delayed melatonin timing and attenuated melatonin levels. In contrast, Escitalopram (0.24 mg/kg per day, over a 60-day period) elevated baseline melatonin concentrations without obvious alteration of the underlying onset of melatonin secretion at the dosage evaluated. A behaviorally delayed sleep-wake time (i.e., 5 hours for 10 days) resulted in a corresponding delay in melatonin onset consistent with entrainment to the altered bedtime. Exogenous melatonin (0.06 mg/kg) produced supraphysiologic concentrations, which precluded interpretation of the endogenous circadian phase. These findings demonstrate that contextual influence on at-home DLMO assessment may differ substantially in effect magnitude at the individual level of analysis. Discussion focuses on the distinction between higher-impact, first-order threats and low-negligible-impact “second-order” threats to at-home DLMO measurement validity.

## INTRODUCTION

A core tenet of circadian medicine is that biological timing plays a central role in the regulation and maintenance of health and treatment of disease [1–3]. Dim Light Melatonin Onset (DLMO) is a physiological signature for evaluating internal clock timing and the onset of the biological night [4–7]. The clinical integration of DLMO assessment has historically been limited, and reviews [4, 6, 8–10] suggest that some of the barriers to implementation have been logistic, and others, economic. Until relatively recently, DLMO assessment was largely restricted to specialized clinical research settings, requiring several hours of supervised sampling under controlled dim-light conditions, dedicated personnel, and either cannulated venous sampling or repeated salivary collection. Methodological advances have now enabled DLMO assessments in home environments using self-collected saliva sampling protocols [9, 11–13].

One concern is that at-home assessments introduce contextual factors that may contribute to systematic or unsystematic measurement error in circadian phase estimation (such as food intake, alcohol, caffeine, physical activity, sleep timing, ambient light exposure, medications, and supplements) [9, 14]. The primary advantages of at-home assessments are accessibility, ease-of-use, and low patient burden. However, if patients are required to exert strict control over too many potentially confounding contextual factors, the convenience and accessibility of this approach may be diminished, thereby reducing its translational and clinical value. Importantly, most recommendations regarding these potential confounders are based on evidence from between-group experimental studies that quantify average effects at the population level, providing limited insight into the magnitude, reproducibility, and clinical relevance of these influences within an individual. Although some of the reported effect sizes are modest-to-large [15–17], many are reported as statistically significant but have effect sizes that are comparatively small [18–20]. Consequently, it remains unclear whether these effects are consistently observable within individuals or whether, in some cases, they are negligible relative to an individual’s normal biological variability.

An issue worth further consideration is related to the ‘magnitude of effect’ of a potential contextual factor’s influence on the interpretation of the DLMO assessment. A hypothetical contextual factor may indeed have a reliable influence on a single-time point measurement of melatonin concentration, but would the consequence of that effect be to change the clinical interpretation of the multiple-sample DLMO profile?

In our previous work, we characterized between-person variation in at-home, saliva-based DLMO assessments collected from 553 sleep clinic patients [21]. DLMO profiles were reliably classified into circadian phase endotypes based on melatonin onset timing, melatonin concentration, and inter-sample variation [21]. Six endotype profiles were described: Within the predicted onset window (POW), delayed melatonin onset, advanced melatonin onset, hypermelatoninemia, hypomelatoninemia, and irregular/multipeak patterns. It is possible that some contextual factors previously identified as statistically significant influences on DLMO metrics may have comparatively minimal practical consequences for endotype interpretation at the intra-individual level. If so, it may be possible to distinguish between first-vs. second-order threats to at-home DLMO measurement error and thereby reduce some of the practical burden associated with self-facilitated, at-home circadian phase assessment.

In the present study, we apply a unique approach to begin to explore this possibility. We apply an N-of-1strategy to complement existing between-person circadian phase research. We constructed a novel and dense intra-individual dataset consisting of 32 repeated at-home salivary DLMO assessments reflecting 247 saliva samples, obtained across four years from a healthy, 35-40-year-old adult male with stable sleep-wake patterns. The baseline series included 10 separate DLMO assessments conducted in the same location and environment on different evenings in consistent dim light. The challenge series involved 23 additional DLMO assessments performed following systematic manipulations of individual contextual factors identified from the literature as having potential confounding effects on melatonin assessment, including; exogenous melatonin administration, ambient and bright-light exposures, blue-blocking glasses [15–17, 22], diazepam [23], behaviorally delayed sleep-wake scheduling [24–26], melatonin-rich foods, magnesium supplementation, caffeine, alcohol, late-evening anaerobic exercise [14, 18–20, 27, 28], and a seasonal photoperiod comparison (winter vs. summer solstice) [29]. We anticipated high intra-individual stability in DLMO timing and endotype assignment across the baseline series. We further anticipated that some contextual manipulations (i.e., bright light exposure, behaviorally altered sleep-wake timing, and endogenous melatonin administration), more than others (i.e., caffeine, food intake, physical activity, or magnesium supplementation), would alter melatonin levels, onset timing, and endotype determination.

## METHODS

### Participant

At the start, the participant was a 35-40 year old adult, Caucasian male volunteer with a self-reported history of stable sleep-wake patterns and only occasional sleep disturbances (< 3 episodes/month). Over the preceding four years, the participant reported no significant sleep-related complaints, chronic medical illnesses, or psychological conditions. Routine medication use was minimal and limited to omeprazole 40 mg/PRN. The participant described rapid adaptation to modest changes in sleep timing, including travel-related jet lag and daylight-saving time transitions, typically within 24 hours and without significant daytime impairment. Typical routine bedtime varied between 9:15 PM to 10 PM, with a routine average bedtime around 9:30 PM. However, variation in routine zeitgebers or bedtime schedule reportedly had minimal impact on sleep initiation or sleep maintenance. Typical sleep latency was reported within 15-30 minutes. The participant identified having an evening chronotype preference, favoring later bedtime and wake times, and reporting greater subjective alertness under delayed sleep schedules. The participant also reported occasional increased sleep latency and poor sleep maintenance during periods of high emotional stress.

### Dim Light Melatonin Onset Assessment Procedure: Baseline Series

On 10 different evenings, 7-or 9-sample salivary Dim Light Melatonin Onset (DLMO) assessments were conducted using a commercially available Clinical Circadian Phase Assessment protocol (Salimetrics, Carlsbad, CA). Assessments for the baseline series were performed in the same environmental location at the participant’s home under controlled and consistent dim light conditions. Ambient illumination ranged from approximately 2 to 8 lux and was measured using a calibrated light meter (Accuracy: ±4% ± 0.5% f.s [0-20,000 Lux]; ±5% ± 10 [20,000-400,000 Lux]) positioned at eye level, within 2-3 inches from the retina during the sampling interval.

During the intra-sample intervals, the participant mainly watched television and remained predominantly sedentary with minimal physical activity. The television was a 50-inch LED display, positioned approximately 12 feet from the participant, with brightness maintained at the lowest setting. Standard sunglasses were worn to minimize direct retinal light exposure. These assessments involved the collection of saliva beginning 5 hours prior to habitual bedtime and continuing until 1 hour after bedtime at approximately 1-hour intervals.

On two occasions, a baseline series of 7 or 9-sample Dim Light Melatonin Offset (DLMOff) was also performed to enable comparison to a condition in the challenge series. These assessments began upon waking (∼5:30 AM) and continued at either 30-minute or 1-hour intervals until all samples were collected.

### Saliva Collection and Determination of Melatonin

At each saliva collection time point, approximately 0.5 mL of whole saliva was collected via the passive drool method into a 2 mL cryogenic vial. Samples were immediately frozen in the participant’s home freezer, then returned to a centralized laboratory under cold-chain transport conditions consistent with previously published protocols [9, 11, 12]. Following receipt at the laboratory, saliva samples were stored frozen at –80C until the day of assay. On the day of analysis, samples were thawed and centrifuged to remove mucins prior to melatonin quantification. Salivary melatonin concentrations were determined in a CLIA laboratory (Salimetrics Clinical Laboratory, State College, PA) using a commercially available melatonin enzyme immunoassay specifically validated for use with saliva (Salimetrics, Cat # 1-3402). The assay has a lower limit of detection of 1.37 pg/ml, a dynamic range from 0.78 to 50 pg/mL, and reported average intra- and inter-assay coefficients of variation of 5.20% and 8.26%, respectively.

### Calculations of Dim Light Melatonin Onset and Assignment to Endotype

DLMO was estimated using a fixed 4 pg/mL melatonin threshold when baselines were below the assay limit of detection, and the “3k method” when baseline concentrations were sufficient, defined as melatonin concentrations exceeding two standard deviations above the mean of baseline samples, in accordance with prior circadian phase assessment recommendations [5, 9, 30, 31]. We assigned DLMO assessments into 3 primary circadian phase profiles based on the timing of DLMO relative to bedtime: Predicted Onset Window DLMO (POW) defined as melatonin onset occurring within 1–3 hours before habitual bedtime, with peak melatonin concentrations exceeding 10 pg/mL; Delayed Melatonin Onset defined as melatonin onset occurring within 1 hour of, at, or after self-reported bedtime; and Advanced Melatonin Onset defined as melatonin onset occurring more than 3 hours before reported bedtime [21, 32, 33].

Two additional phenotype profiles were considered to capture the potential influence of medications that either directly or indirectly (via iatrogenic factors) elevated or attenuated melatonin secretion [23, 34]. Hypermelatoninemia was defined as elevated melatonin levels (>10 pg/mL) without a clear onset pattern, whereas Hypomelatoninemia was defined as little to no consistent rise in melatonin, or melatonin levels that remained consistently below 10 pg/mL for more than 85% of sampling time points.

## RESULTS

### Overview

Results are organized by specific conditions that may be encountered during at-home clinical testing. Under research protocols, these are typically highly controlled. A brief description of each specific method is applied, and figures are presented with baseline comparisons to illustrate intra-individual patterns of melatonin production and circadian phase behavior across repeated at-home salivary DLMO assessments.

### DLMO timing stability across repeated assessments under standardized conditions

Ten baseline, 7-sample DLMO assessments were completed under standardized dim light conditions across approximately 16 months between August 2022 and July 2024. Five DLMO assessments were conducted weekly between August and September 2022, with an additional assessment completed approximately 30 days later. Four subsequent baseline assessments were then completed across a four-month interval beginning in July 2023, with a final assessment completed approximately 1 year later, in July 2024. Each assessment was performed using the same standardized collection procedure described above. Figure 1 presents the averaged baseline melatonin profile derived from 10 assessments.

**Figure 1:**
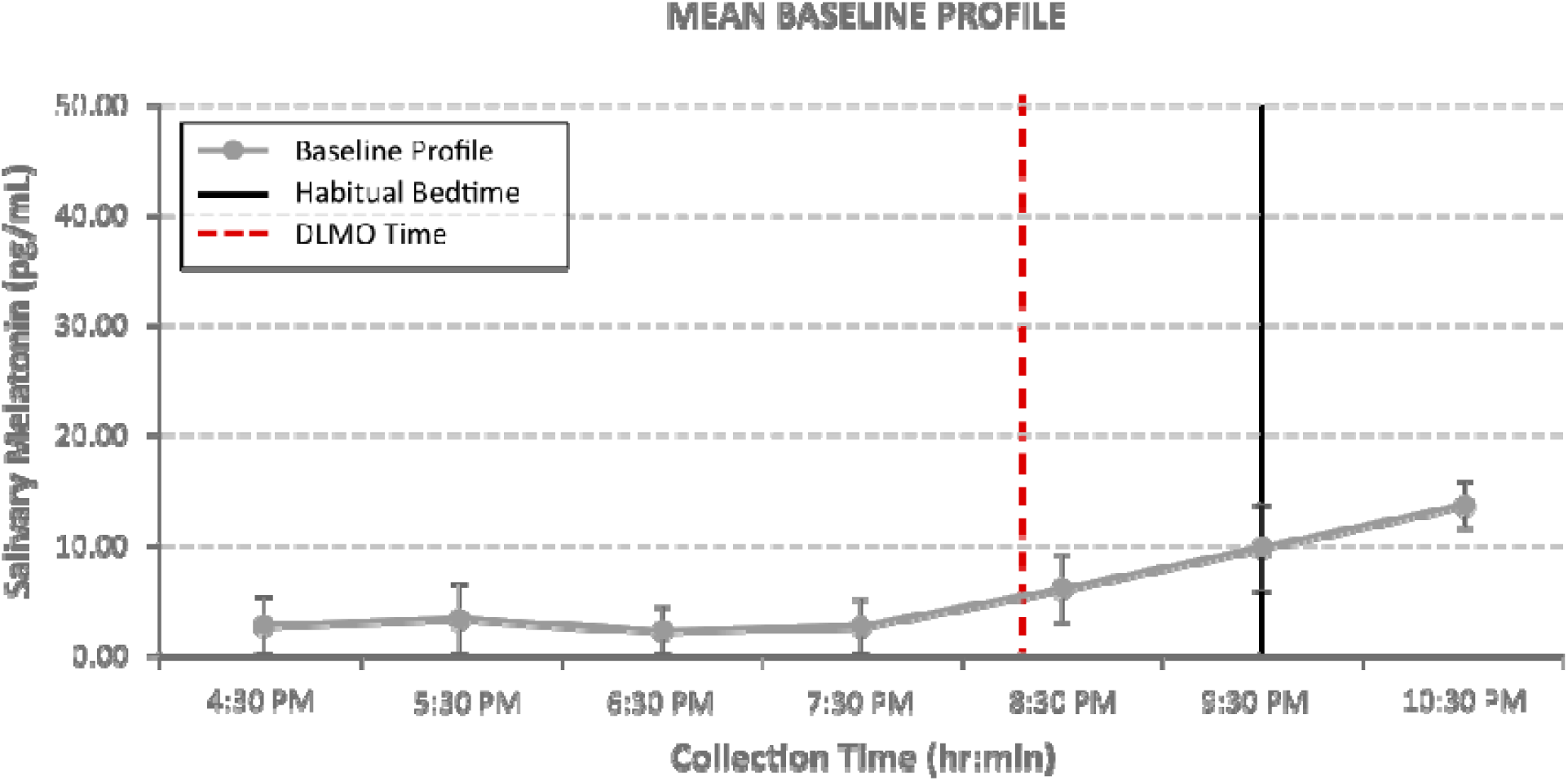
Baseline salivary melatonin profile (n = 10): Mean salivary melatonin (and standard errors) by sampling interval. Red dotted line indicates the average DLMO time. Black vertical line indicates habitual bedtime.

Across the 10 baseline assessments, mean DLMO timing was 8:19 PM (SD = 28 min; SE = 9 min), with individual assessments occurring within a relatively narrow 90-minute window (7:13 PM–8:43 PM). The mean absolute deviation (MAD) from the participant’s long-term mean DLMO was 22 minutes, demonstrating remarkable intra-individual consistency despite assessments spanning weeks, months, and seasons. All baseline assessments were independently classified by two blinded raters according to previously published endotype decision rules, with 100% inter-rater agreement. All assessments were classified within the Predicted Onset Window endotype. Collectively, these findings demonstrate substantial long-term stability in both circadian phase timing and melatonin profile morphology in a well-entrained individual.

### Exogenous melatonin supplementation

A baseline DLMO assessment was performed prior to administration of 5 mg exogenous melatonin (Spring Valley Fast Dissolve, 5 mg) at 7:35 PM (Figure 2). Subsequently, a 9-sample morning melatonin offset was then performed on the following morning, beginning at 5:30 AM until 1:30 PM (Figure 3).

**Figure 2:**
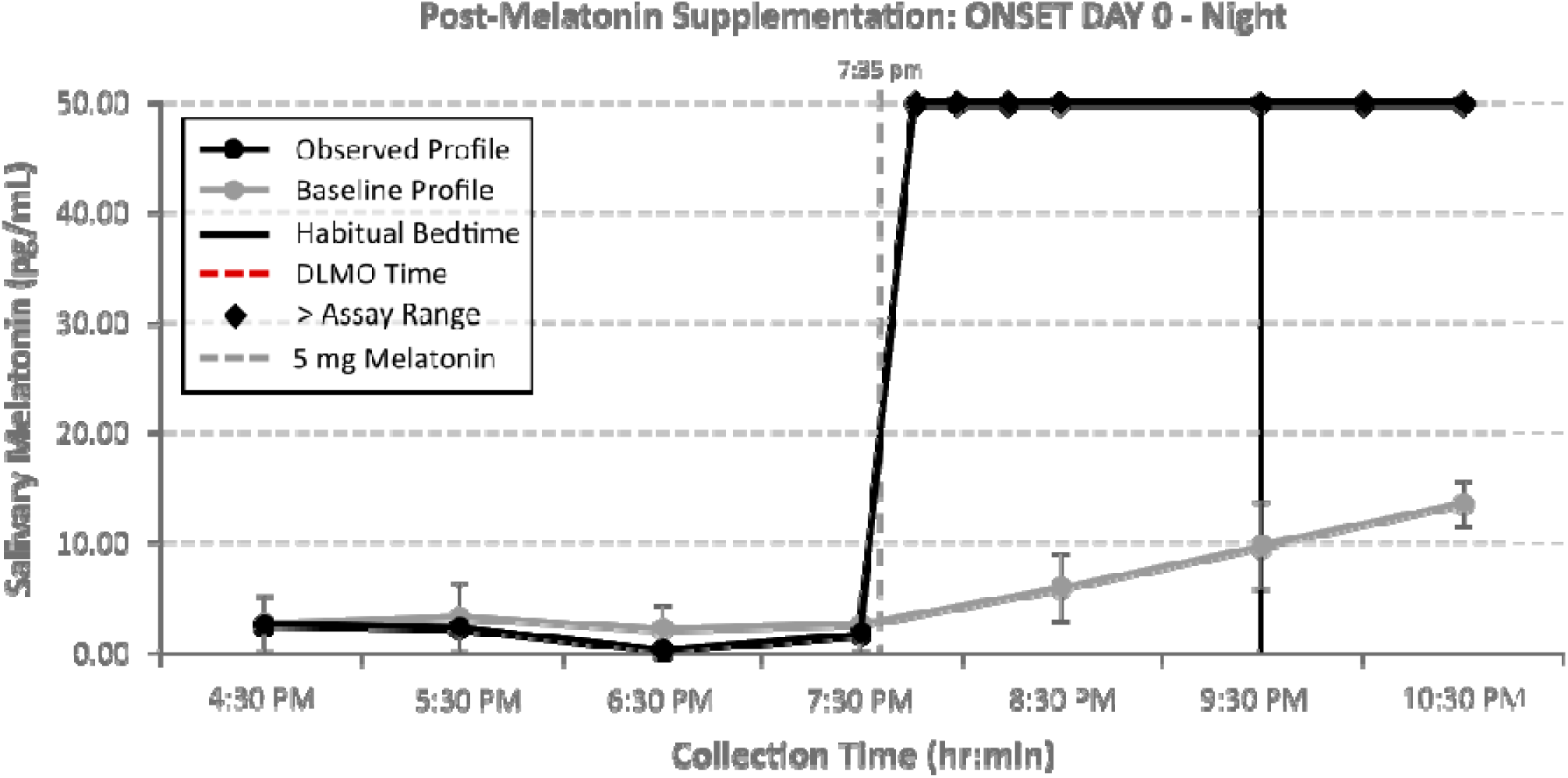
Salivary melatonin concentrations before and after 5 mg exogenous melatonin supplementation. For all subsequent figures, the solid grey line represents the average baseline melatonin profile unless otherwise specified.

**Figure 3:**
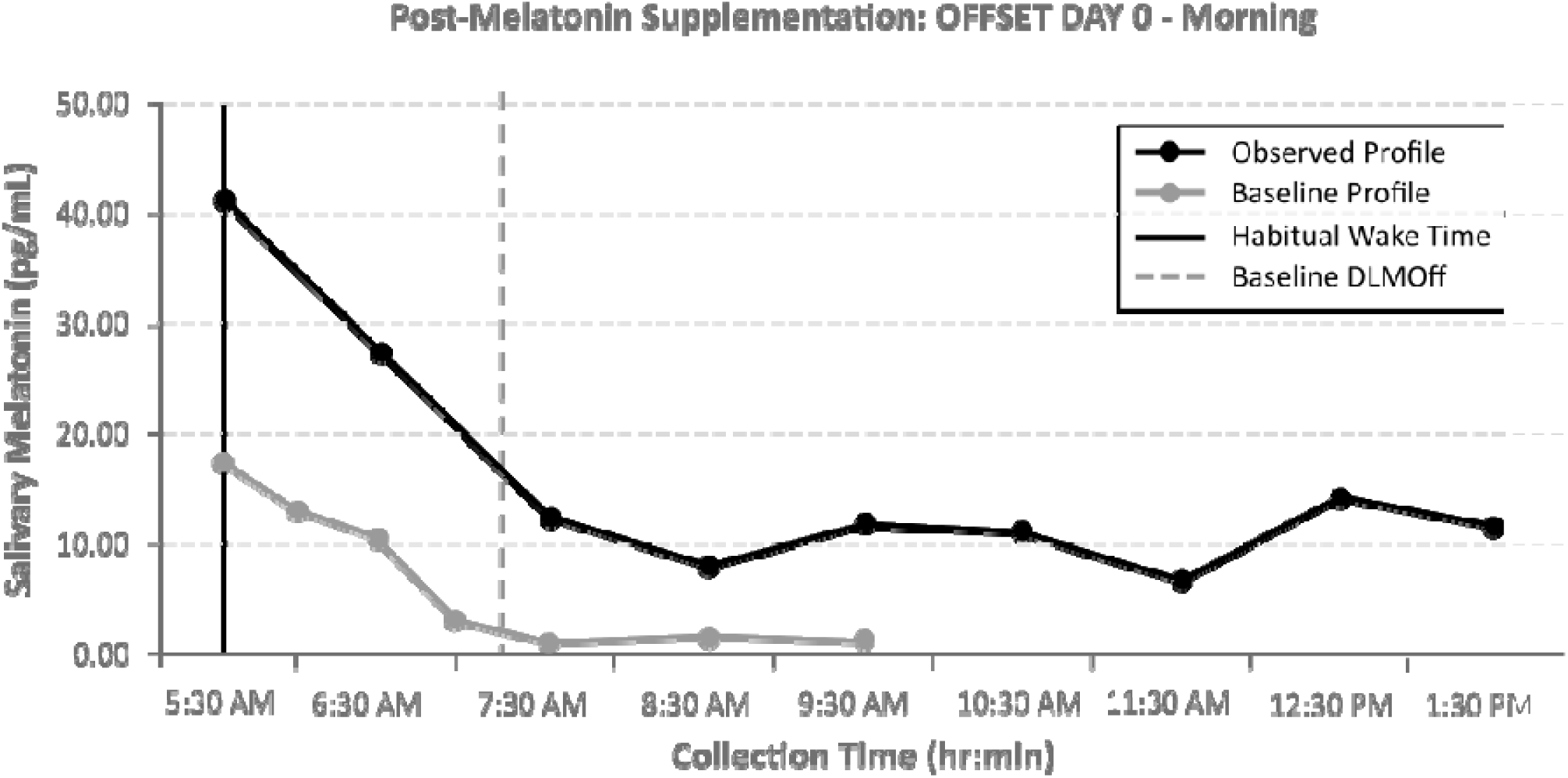
Morning melatonin profile (in dim light) following evening exogenous melatonin supplementation. Baseline profile was established by averaging two Dim Light Melatonin Offset profiles taken pre-supplementation on two different occasions.

**Figure 5:**
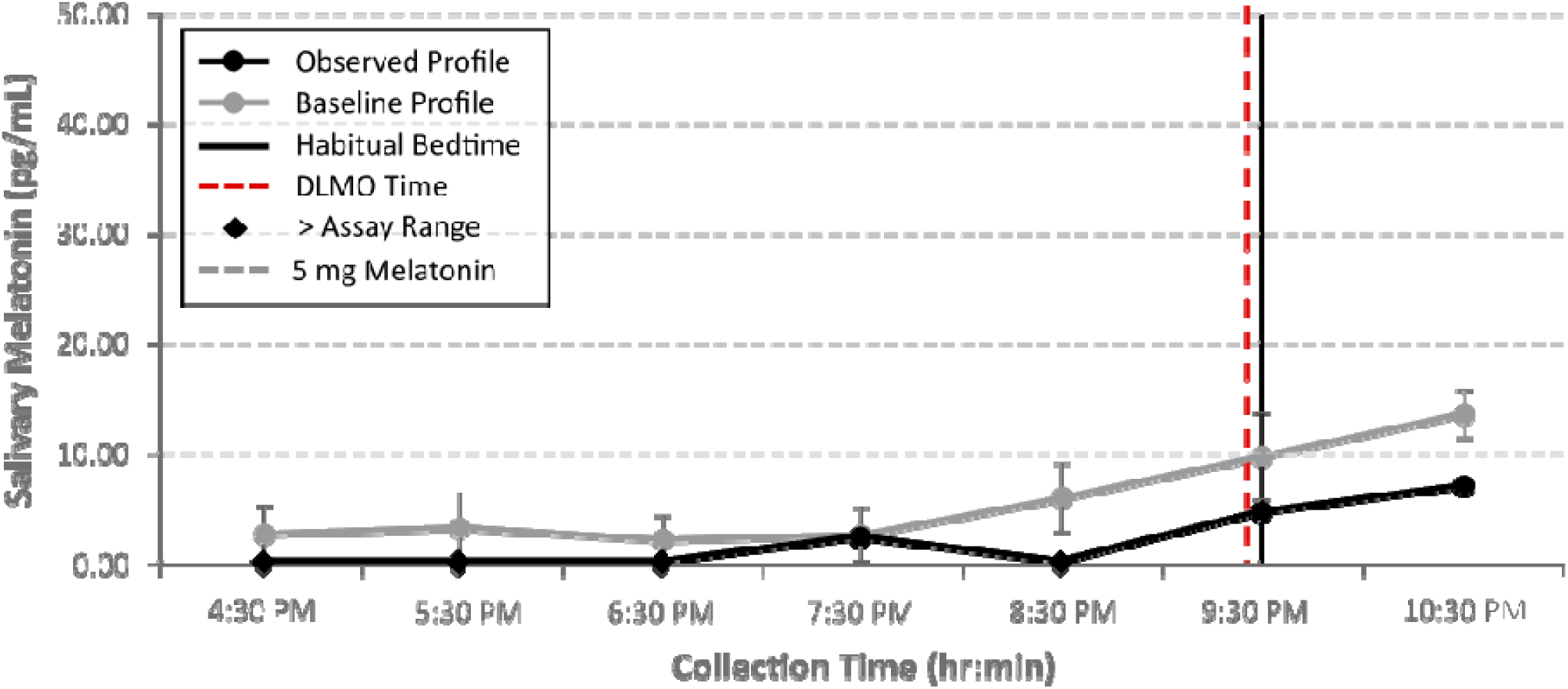
Salivary melatonin levels obtained under unrestricted ambient light conditions.

Melatonin levels exceed the assay detection range within approximately 10 minutes of melatonin supplementation and remained elevated throughout the evening assessment period, precluding interpretation of endogenous circadian phase timing or endotype.

Melatonin concentrations remained elevated the morning following supplementation and gradually declined across the post-wake period, although concentrations remained above typical baseline daytime concentrations during portions of the sampling interval.

### Impact of ambient light exposure on DLMO assessment

A 7-sample DLMO was performed under unrestricted environmental lighting conditions without dim light controls. The preceding ambient illumination ranged from approximately 54 lux (pre-sunset) to 17 lux (post-sunset with interior lighting on). Sunset was estimated at 8:01 PM.

Relative to baseline conditions, melatonin onset appeared delayed and overall melatonin concentrations were reduced during unrestricted light exposure compared to baseline.

### Impact of blue-light blocking glasses during unrestricted ambient light exposure

A 7-sample DLMO assessment (see Figure 6) was repeated under unrestricted household lighting conditions (same as above) while wearing commercially available <u>ZICOTO Blue Light</u> <u>Blocking Glasses</u> (Amazon). Ambient interior Lux this evening ranged between approximately 33 lux (pre-sunset) to 17 lux (post-sunset with interior lighting on). Sunset was estimated at 6:35 PM.

**Figure 6:**
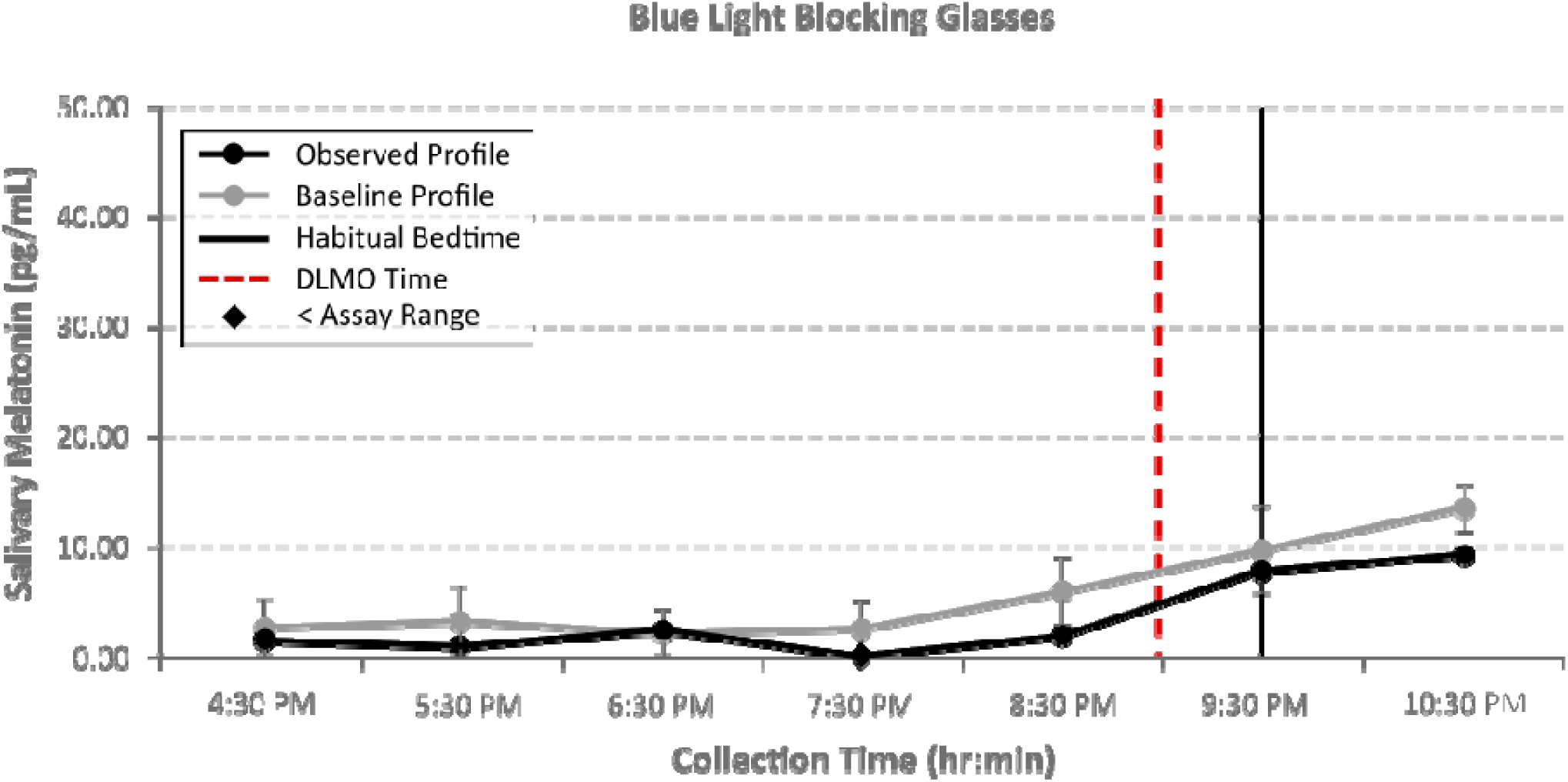
Salivary melatonin profile using blue-blocker glasses during sample collection and no dim light control.

Compared with unrestricted ambient light exposure (baseline profile, grey line), melatonin onset remained detectable, and overall profile morphology appeared similar to baseline conditions, although peak concentrations remained somewhat attenuated relative to standardized dim light controlled assessments.

### Impact of continuous, high-intensity bright light exposure

Multiple DLMO assessments were performed under controlled bright light exposure conditions (∼1500 lux). Two 750 Lux, 5600k temperature-calibrated lights were placed approximately 2 feet from the subject at ∼60° and ∼120° adjacent to the direct line of sight. Light output as measured by a light meter placed at eye level, was recorded at approximately 1500 lux.

Continuous bright-light exposure initiated prior to the expected DLMO resulted in marked suppression of melatonin production throughout the assessment interval (Figure 7).

**Figure 7:**
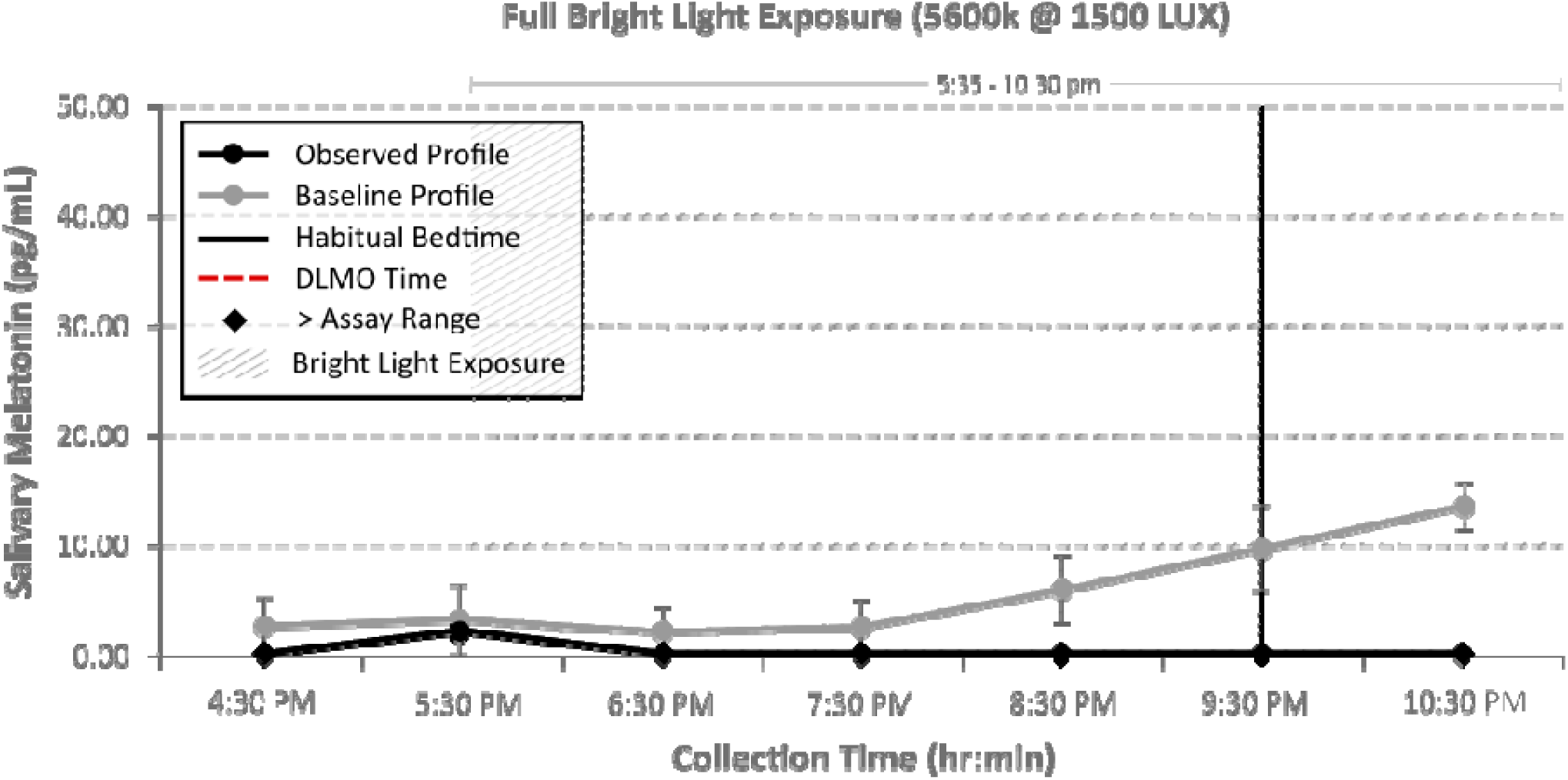
Salivary melatonin profile during continuous pre-onset bright light exposure (∼1500 lux).

Additional assessments evaluated bright-light exposure initiated after DLMO onset. Under these conditions, melatonin concentrations demonstrated significant declines following continuous exposure to high-intensity light. Exposure to 5600K light produced a steeper decline in melatonin concentrations than exposure to 3200K light of comparable intensity (Figure 8). Brief 5-minute bright-light exposure after onset produced only transient reductions in melatonin concentrations, with subsequent recovery toward baseline levels during later sampling intervals (Figure 9). In a prolonged bright-light exposure condition extending until 9:30 PM, melatonin concentrations remained markedly suppressed initially but subsequently demonstrated gradual recovery following lights-out (Figure 10). Finally, DLMO timing was assessed during the summer and winter solstices to determine whether seasonal differences in photoperiod duration were associated with measurable shifts in circadian phase timing (Figure 11).

**Figure 8:**
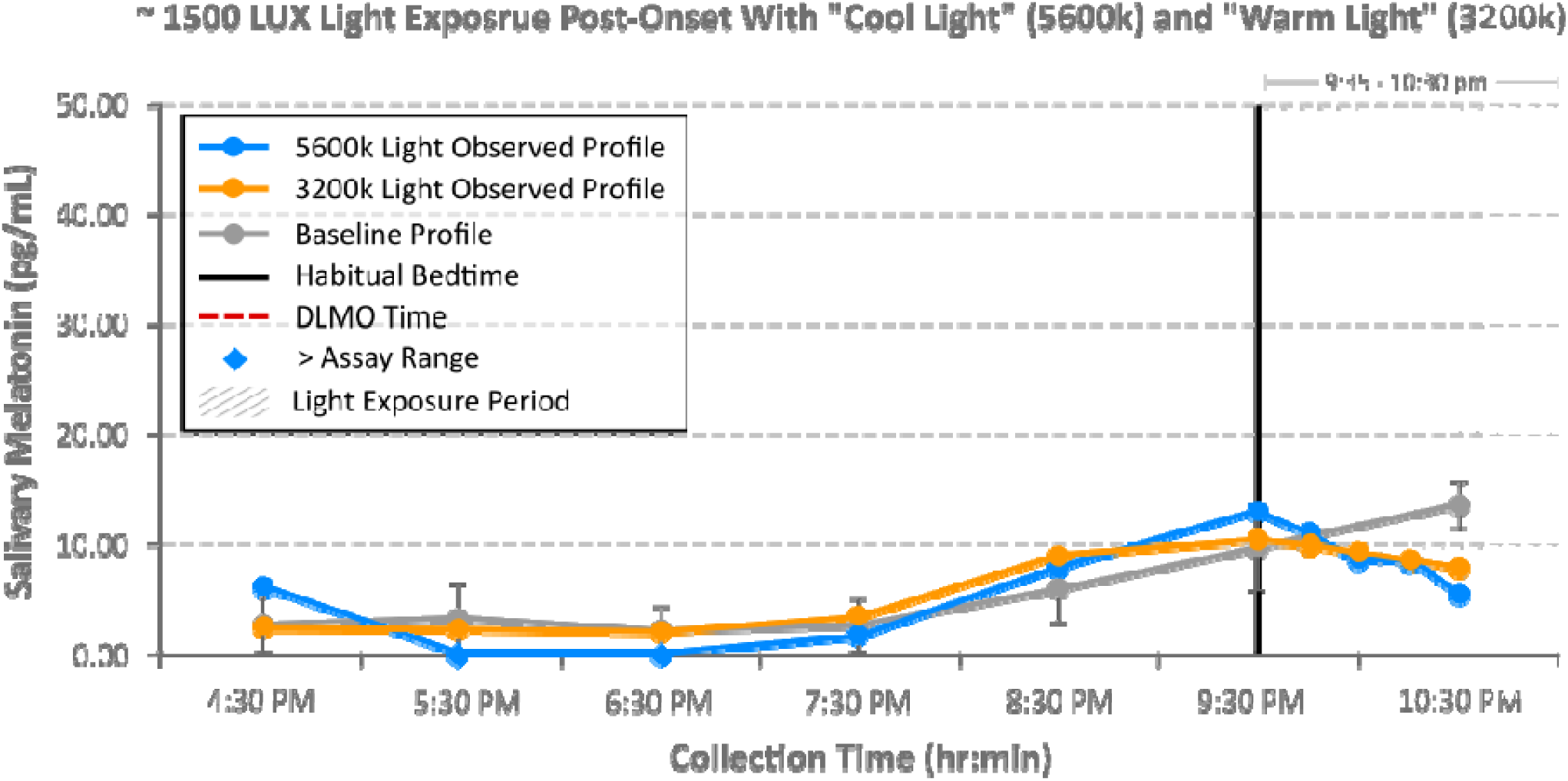
Comparison of salivary melatonin profiles during post-onset bright light exposure (∼1500 lux) at 5600k and 3200k light vs baseline.

**Figure 9:**
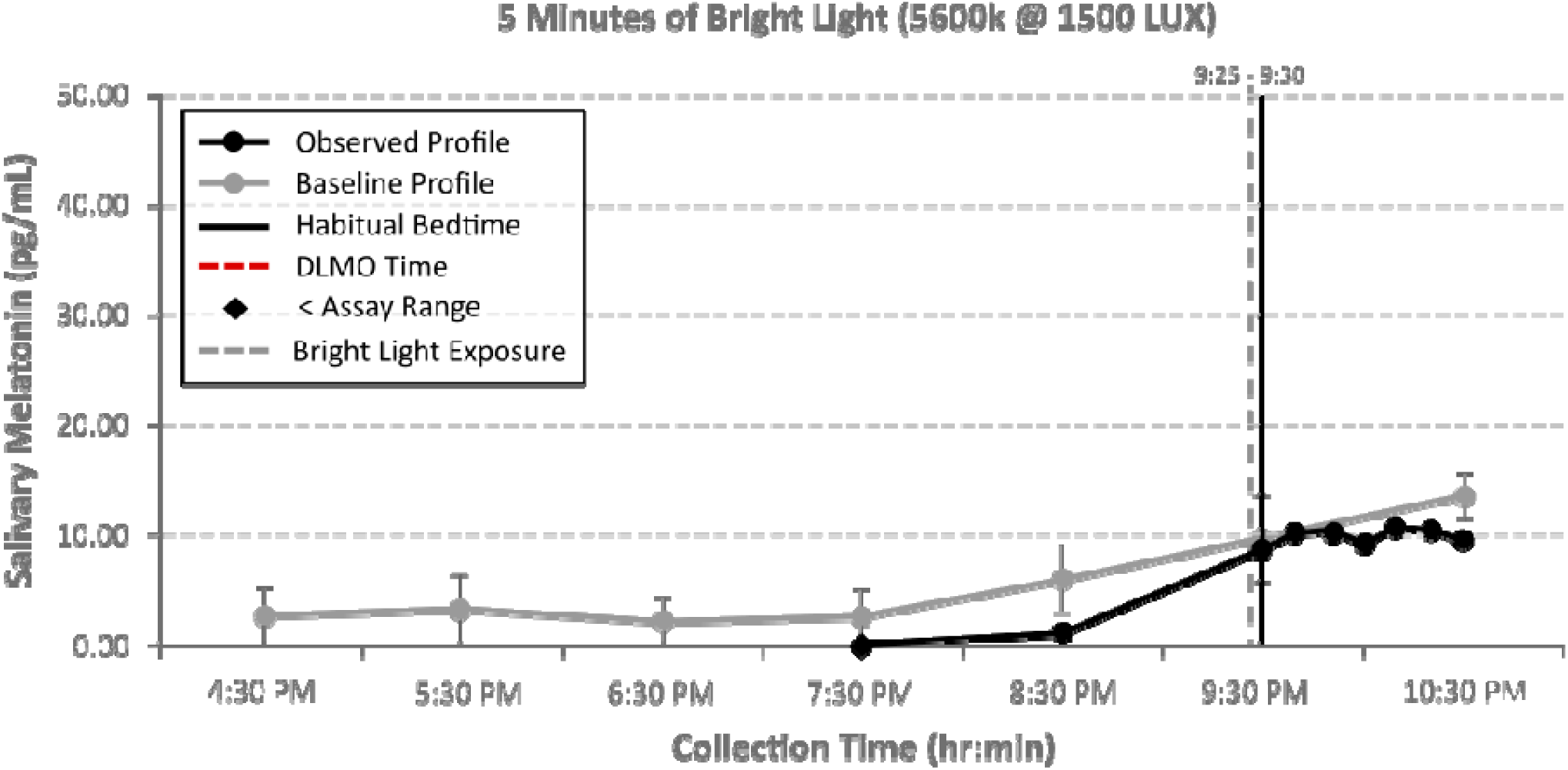
Salivary melatonin profile after 5 minutes of bright light exposure (5600k @ ∼1500 lux) post-onset.

**Figure 10:**
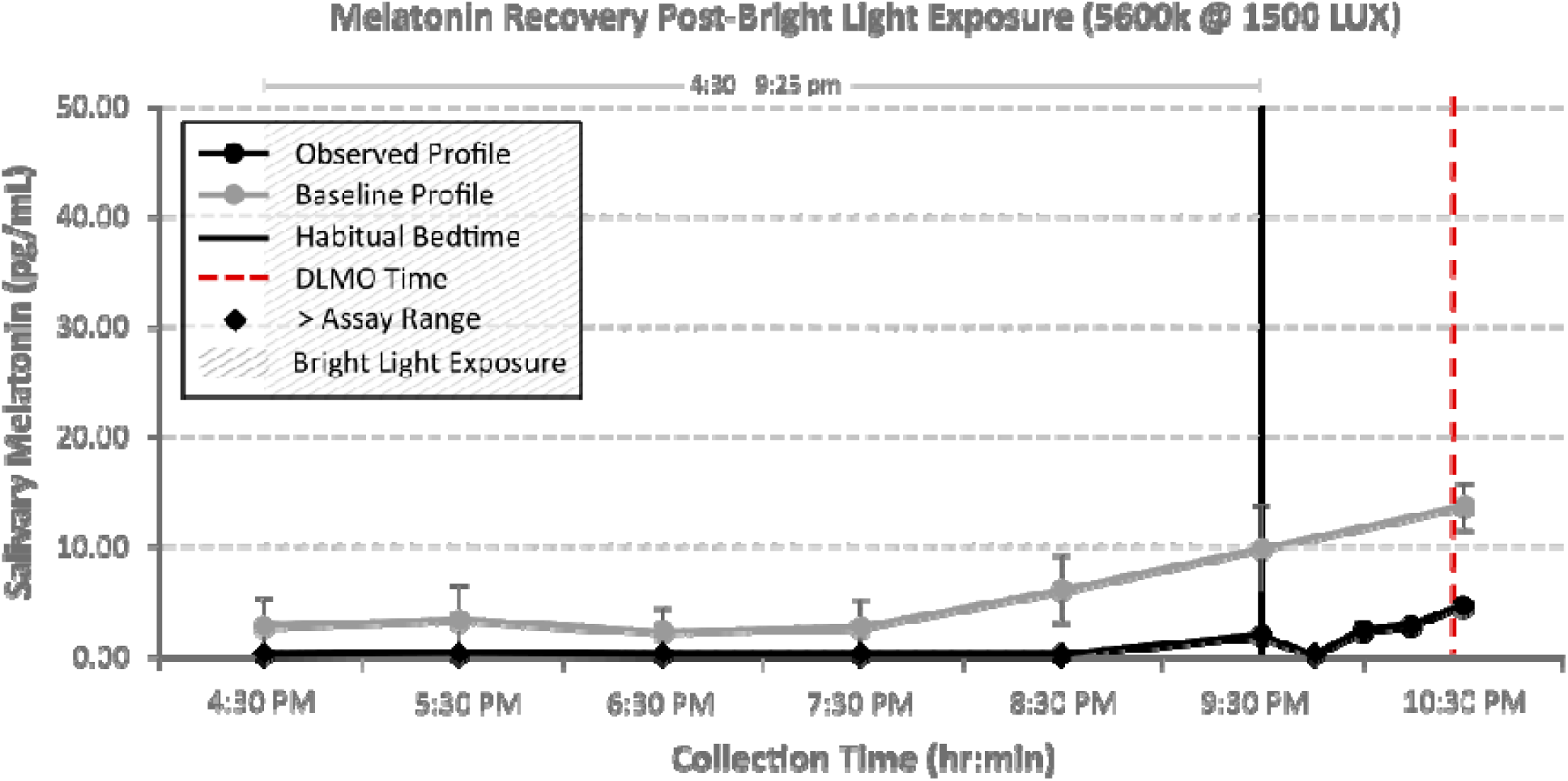
Salivary melatonin profile recovery after continuous bright light exposure (5600k @ ∼1500 lux) post-onset.

**Figure 11:**
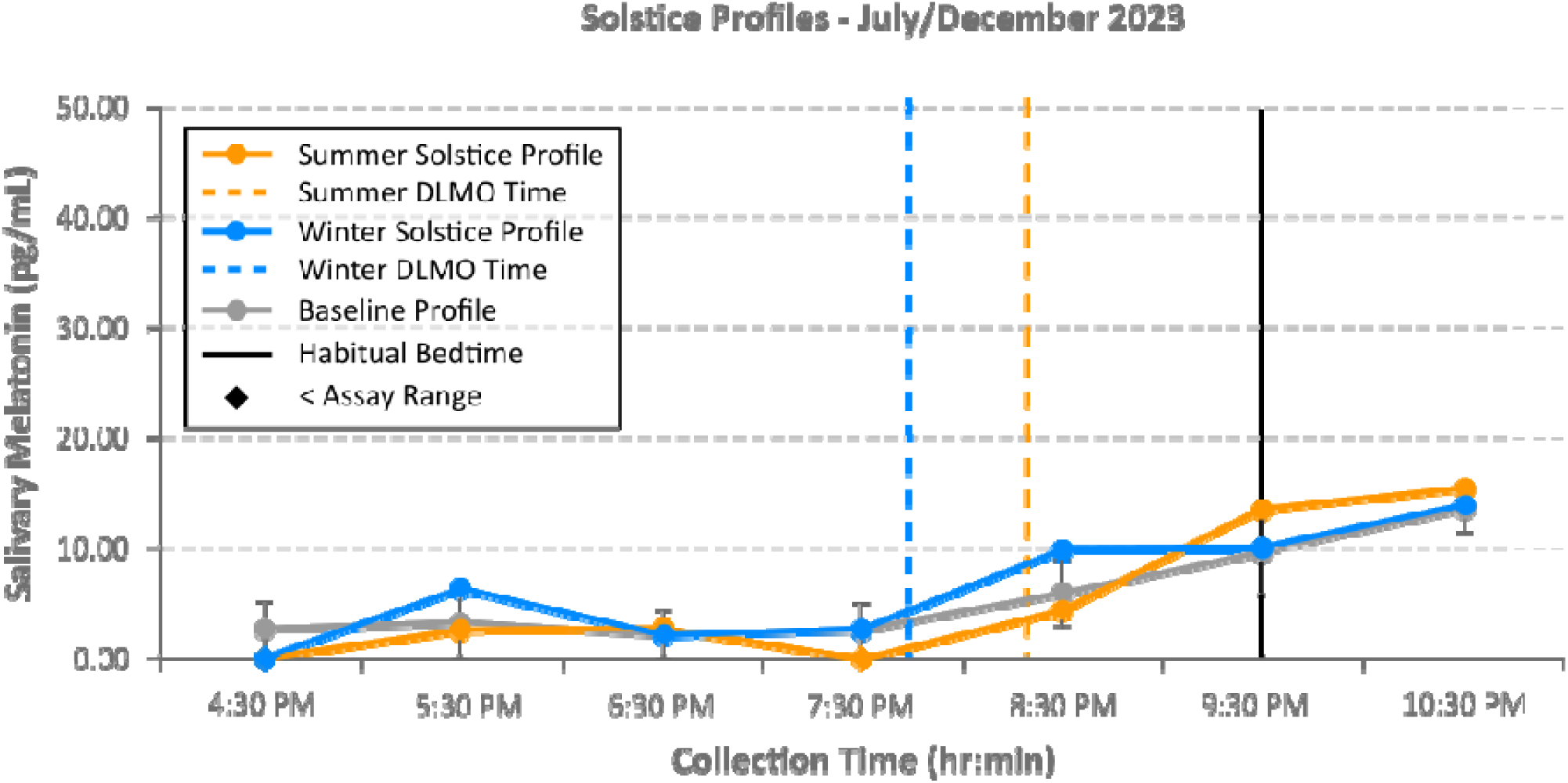
Seasonal variation in salivary melatonin profiles during winter and summer solstices.

### Impact of a benzodiazepine and Selective Serotonin Reuptake Inhibitors (SSRIs)

Diazepam exposure (10 mg daily for 5 days, with an additional 10 mg dose administered approximately 1 hour prior to onset was associated with lower melatonin concentrations and later apparent onset timing relative to baseline profiles (Figure 12).

**Figure 12:**
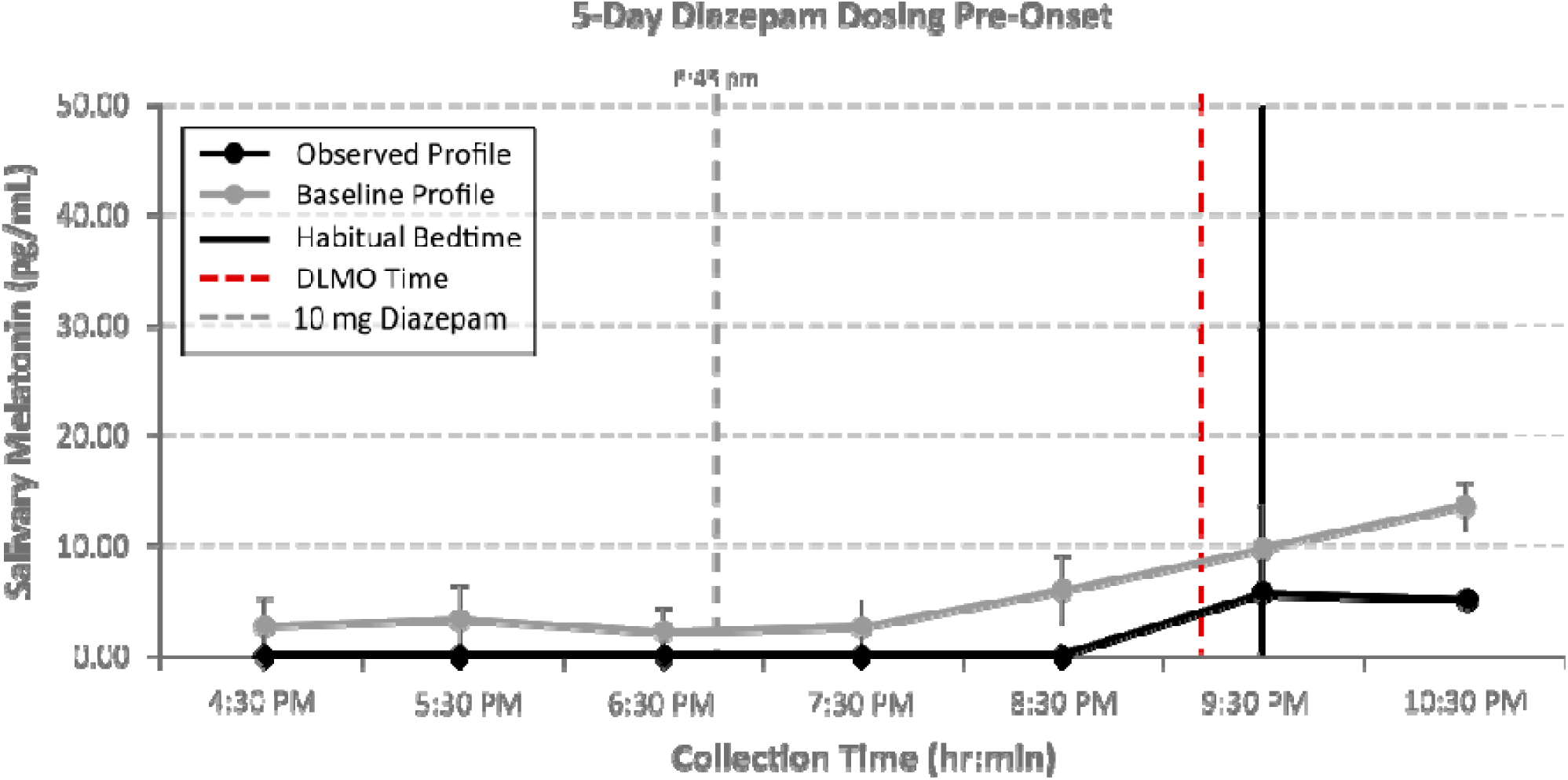
Salivary melatonin profile following diazepam exposure (10 mg daily for 5 days with an additional 10 mg dose administered approximately 1 hour prior to expected DLMO onset).

Escitalopram (SSRI) exposure (20 mg daily for 60 days) was associated with modestly elevated baseline melatonin concentrations relative to baseline assessments, although overall onset timing and profile waveform remained visually comparable to the participant’s established baseline profile (Figure 13). The apparent delay in calculated DLMO was attributable to the elevated melatonin baseline concentrations rather than a discernible phase shift.

**Figure 13:**
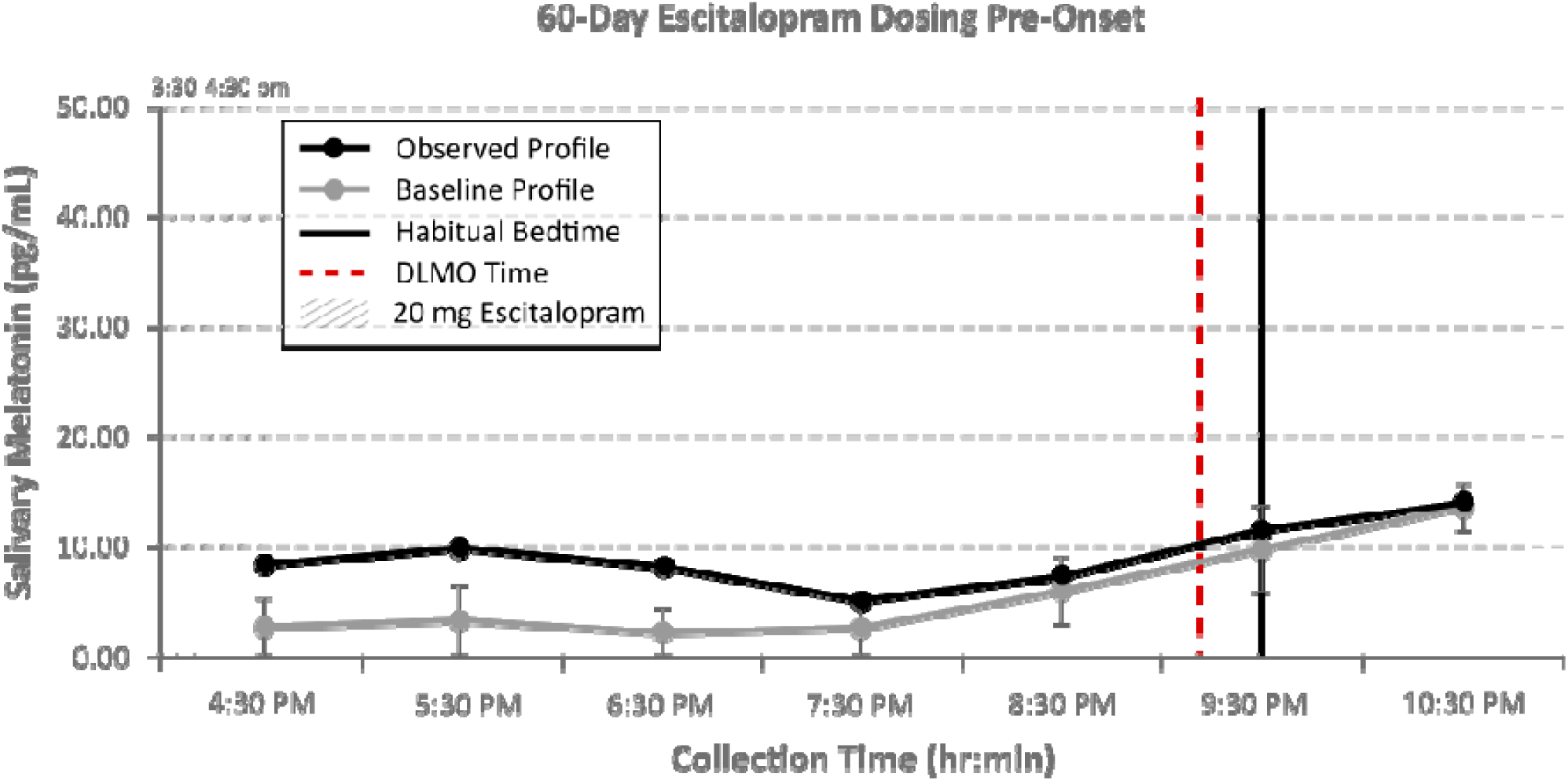
Salivary melatonin profile following escitalopram (SSRI) exposure (20 mg daily for 60 days).

### Impact of behaviorally delayed sleep timing

DLMO assessments were performed before and after a 10-day period during which bedtime was delayed by approximately 5 hours relative to the participant’s habitual schedule. This experiment was designed to evaluate circadian adaptability when a 5-hour bedtime delay was routinely instituted from a normal, routine bedtime (Figure 14).

**Figure 14:**
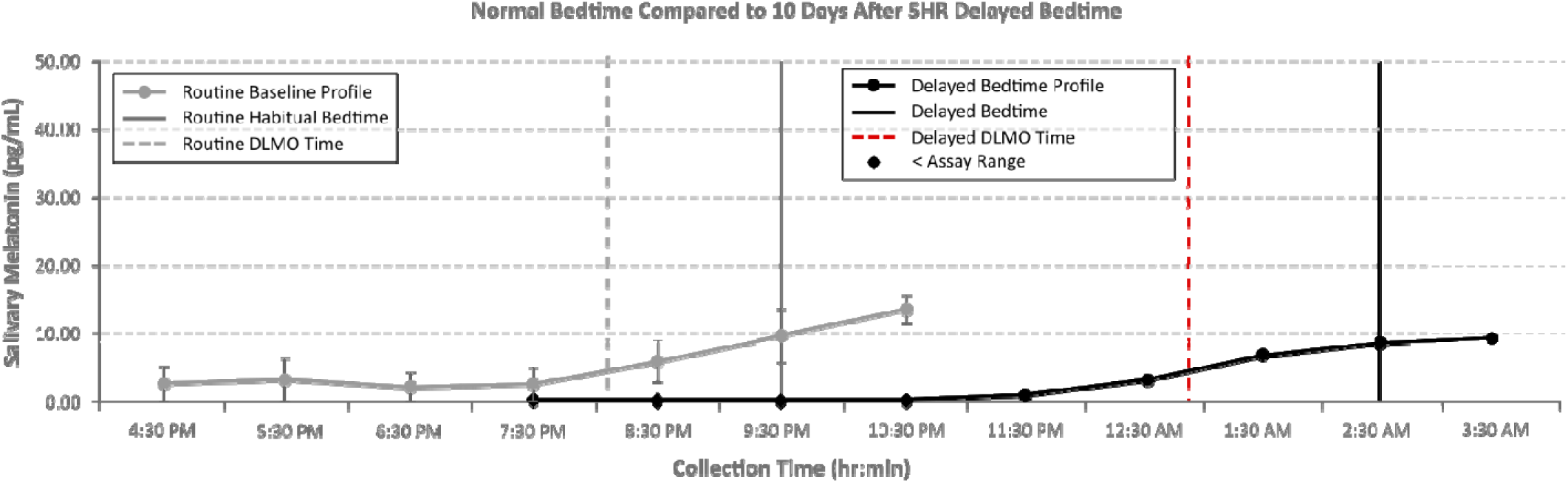
Salivary melatonin profiles before and after a 10-day, 5-hour behavioral sleep schedule delay relative to the participant’s typical routine.

Following the delayed sleep schedule interval, DLMO timing shifted later in approximate alignment with the delayed bedtime schedule, preserving overall profile waveform and peak melatonin concentrations relative to baseline conditions. These observations are consistent with adaptive circadian phase delay in response to sustained behavioral bedtime modification.

To further examine transient behaviorally delayed sleep timing, an additional DLMO assessment was performed following a 5-hour delay in habitual bedtime maintained over two consecutive weekend evenings after re-establishing the participant’s baseline sleep schedule and profile. The assessment was designed to characterize short-term changes in circadian phase associated with temporary behavioral schedule delay to simulate a common pattern of social jet lag. (Figure 15).

**Figure 15:**
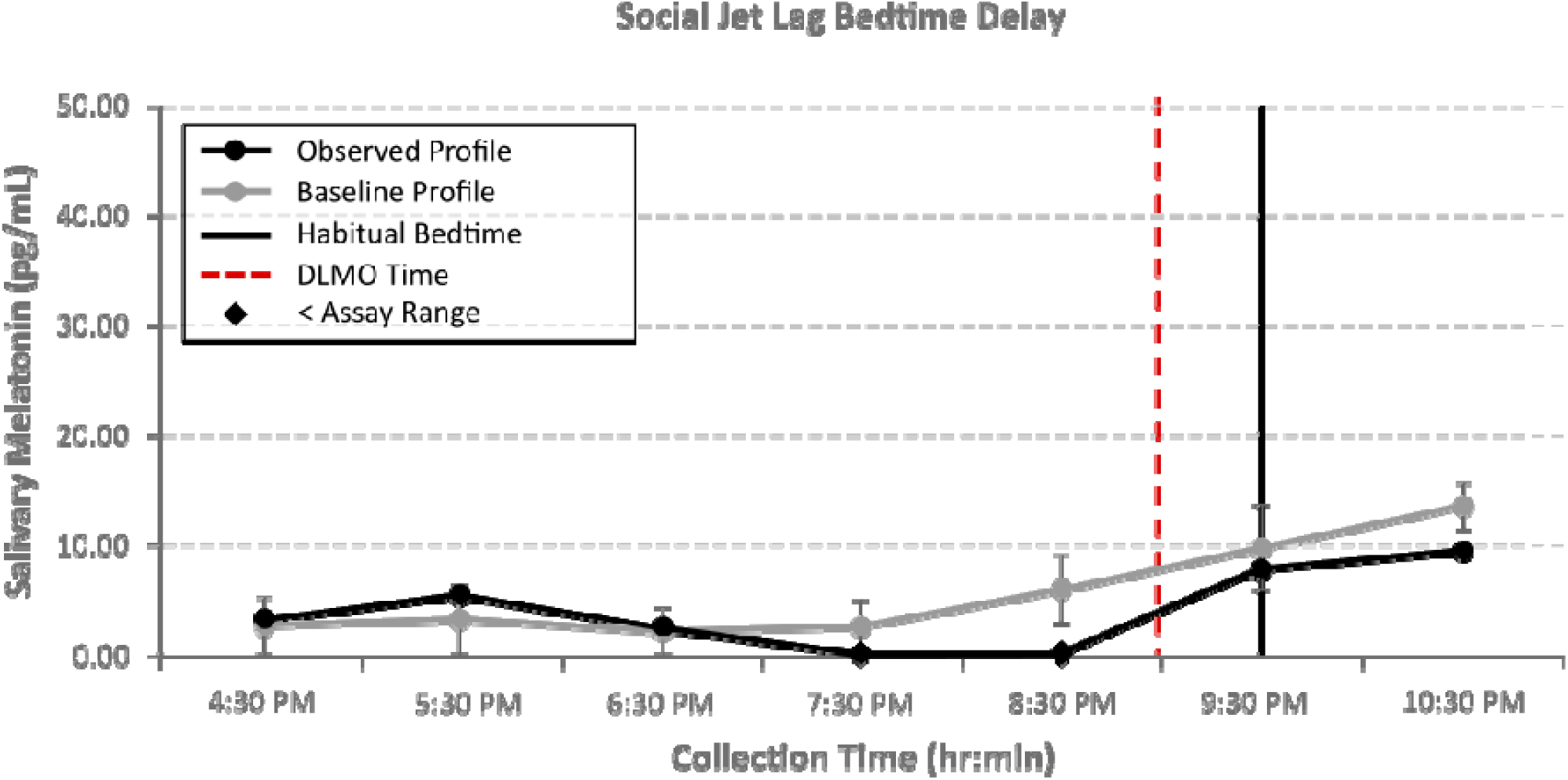
Salivary melatonin profile after a 2-day, 5-hour behavioral sleep schedule delay relative to the participant’s typical routine.

### Impact of commonly cited contextual influences

Additional DLMO assessments examined the effects of food intake, including melatonin-rich foods, magnesium glycinate supplementation (4 days at 300 mg), caffeine intake (300 mg), alcohol consumption (6oz of 80-proof rum), and 45 minutes of anaerobic exercise during the collection window (post-onset) on DLMO timing and peak concentrations. Although minor fluctuations in individual sampling points were observed across some conditions, these differences were generally within the range of baseline intra-individual variability.

During baseline collections, the following meals were consumed:

- Spaghetti and Meatballs with Marinara Sauce (x3) (beef, pasta, tomatoes, onions, cheese)
- Jersey Mike’s Italian Sub (deli meat, provolone, lettuce, onions, tomatoes, black olives)
- Huevos Rancheros Tacos (eggs, avocado, tomatoes, onions, feta, spinach)
- Veggie Skillet Meal (black beans, potatoes, onions, green peppers, tomatoes, mushrooms, cream cheese)
- Chili (tomatoes, kidney beans, pinto beans, onions, green peppers, beef)
- Stuffed Cabbage (cabbage, rice, tomatoes, onions, beef)
- Veggie Burger /w Steamed Corn (Beyond Meat Burger, tomatoes, onions, cheese)
- Cajun Chicken Alfredo (alfredo sauce, parmesan cheese, chicken, pasta, red peppers) Additionally, two melatonin-rich meals were consumed;

#### Melatonin Rich Meal 1

Black Cherry Char Siu Pork Stir-Fry with white rice, raisins, cashews, almonds, cabbage, green pepper, onions, and collard greens, and an 8oz glass of tart cherry juice (Lakewood Organic Tart Cherry Concentrate Juice), (Figure 16).

**Figure 16:**
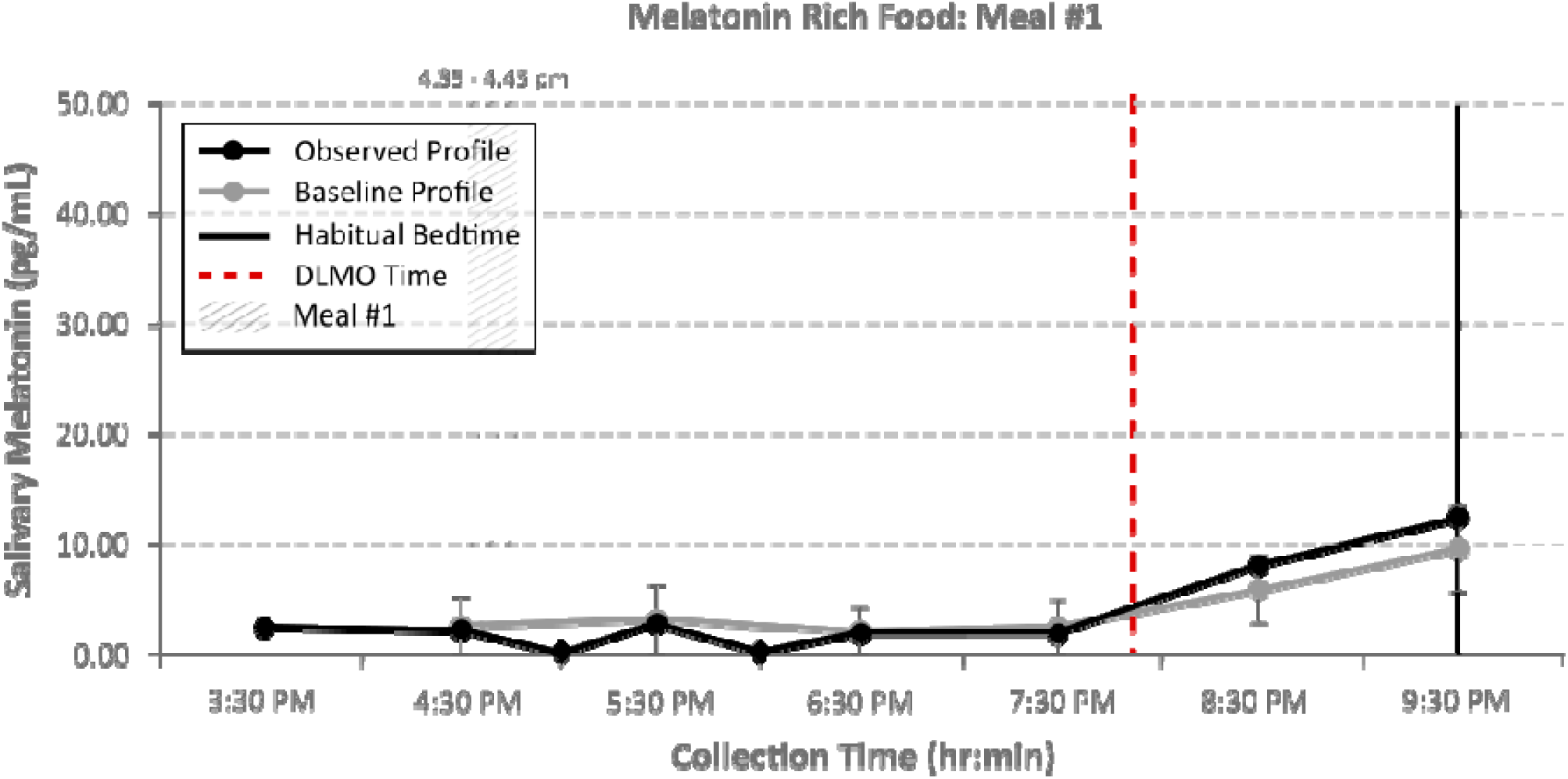
Salivary melatonin profile following consumption of a melatonin-rich meal #1.

#### Melatonin Rich Meal 2

8 oz of organic dark sweet cherries (frozen), 1oz pistachios, 1 slice of pumpkin pie, with a 12oz glass of tart cherry juice (Knudsen Family, Organic). Sampling every 15 minutes for 1 hour-post consumption, and then 30 minutes later to more closely monitor baseline levels (Figure 17).

**Figure 17:**
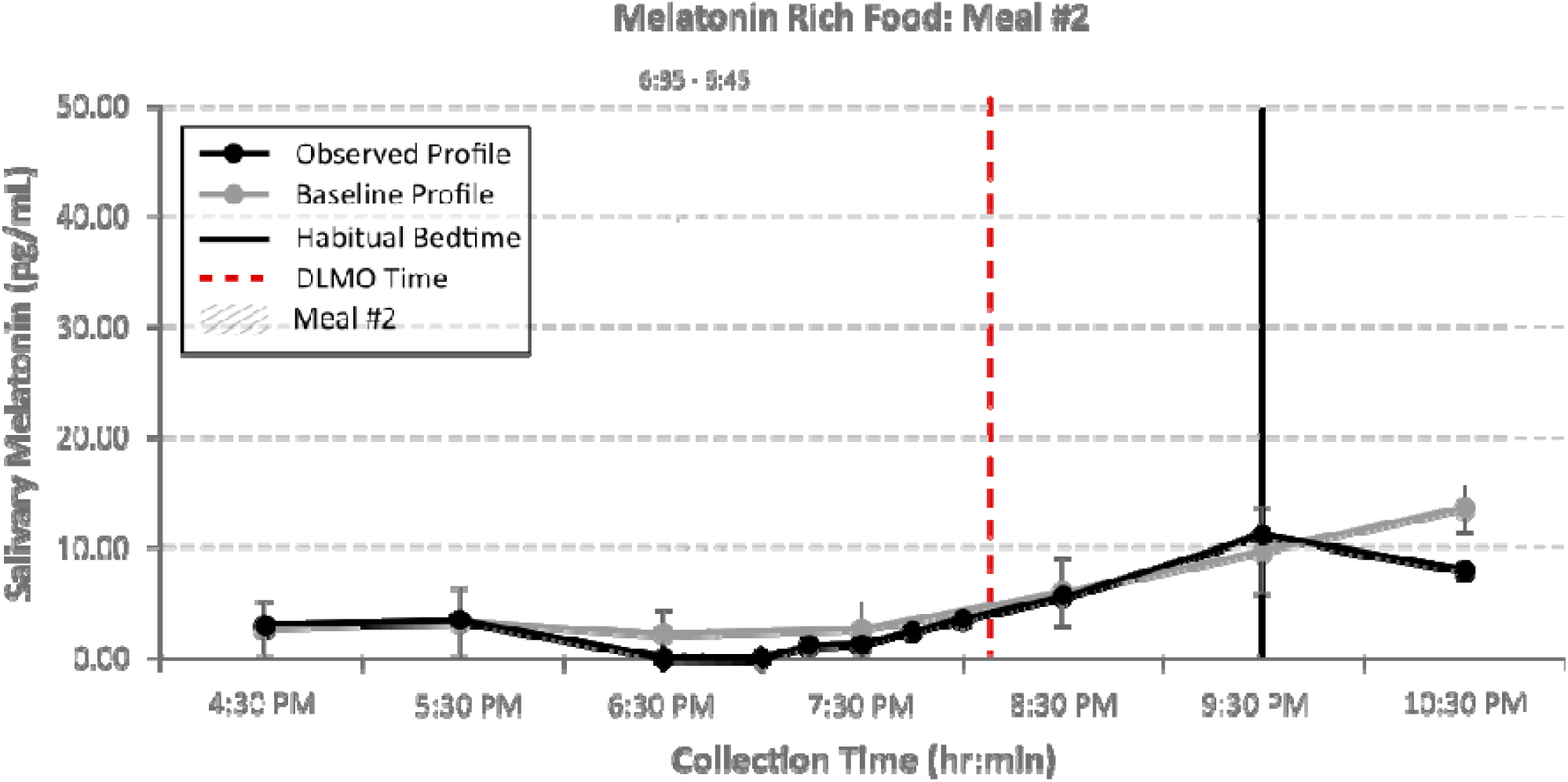
Salivary melatonin profile following consumption of a melatonin-rich meal #2.

Sampling intervals in these profiles were increased to account for the relatively short half-life and rapid metabolism of melatonin.

Across the initial 12 meal conditions, salivary melatonin concentrations remained generally comparable to baseline profiles, with no substantial elevations observed following food intake. Although isolated post-onset samples were modestly elevated relative to baseline means, values remained within the range of baseline intra-individual variability. Meal timing also did not appear to meaningfully alter subsequent DLMO timing under the conditions examined.

## DISCUSSION

If salivary melatonin profiling is to serve as a practical clinical biomarker of circadian phase, it is important to understand the biological and methodological factors that influence DLMO measurements collected under real-world conditions. While most published work has focused on between-subject comparisons, this dense longitudinal N-of-1 dataset provides a complementary perspective by characterizing within-subject stability across repeated assessments obtained over four years.

Under standardized dim-light conditions, DLMO timing and melatonin waveform morphology remained remarkably consistent within the individual despite repeated behavioral, pharmacological, and environmental perturbations. Following temporary circadian disruption, endogenous phase rapidly re-established its baseline timing, suggesting a high degree of circadian resilience under stable entrainment. Onset timing consistently occurred within a relatively narrow clock-time window, and peak melatonin concentrations remained highly reproducible across assessments performed over weeks, months, and seasons [13, 32, 35]. These observations suggest that short-term variability frequently attributed to DLMO may reflect changes in environmental conditions, protocol adherence, or analytical variability more often than instability of the underlying circadian pacemaker itself.

The present findings also contextualize the likely translational relevance of several commonly discussed DLMO confounders. Under the conditions examined, a number of everyday contextual factors commonly treated as potential threats to DLMO interpretation in group-average research, including food intake, caffeine, alcohol, magnesium supplementation, and late-evening anaerobic exercise, produced relatively limited deviations from this participant’s established baseline bandwidth. These contextual factors did not obviously shift the overall profile morphology outside the individual’s baseline bandwidth and did not appear to alter circadian endotype classification or overall DLMO timing. These observations illustrate an important translational possibility: contextual variables capable of producing statistically detectable effects at the group level may not necessarily produce clinically meaningful disruption in circadian phase interpretation for a given individual, particularly when DLMO is interpreted relative to repeated intra-individual baseline assessments [14, 18–20, 27, 28].

Interestingly, diazepam and escitalopram produced opposite effects on melatonin concentrations while exhibiting little evidence of a corresponding change in underlying circadian phase. Diazepam was associated with lower melatonin concentrations and an apparently later DLMO, whereas escitalopram produced modestly elevated baseline melatonin concentrations that similarly resulted in a later calculated DLMO despite preservation of the overall waveform morphology. These observations suggest that pharmacologic interventions may influence melatonin secretion independently of the timing of the circadian pacemaker. Consequently, medications known or suspected of influencing melatonin production, metabolism, or clearance may themselves represent clinically meaningful observations. Accordingly, DLMO should be interpreted within the broader clinical context and in conjunction with the complete melatonin profile, which may provide insight beyond circadian phase alone.

As expected, long-duration bright light exposure during the pre-onset interval represented the largest first-order threat to DLMO interpretability observed in this study. Continuous bright-light exposure markedly suppressed melatonin production and prevented estimation of a representative endogenous circadian phase. Bright-light exposure post-onset also produced rapid declines in melatonin concentrations that did not fully recover within one hour, suggesting that inadvertent post-onset light exposure may continue to influence late-evening melatonin dynamics even after onset timing has already been captured. Collectively, these observations underscore the practical importance of light control during clinical DLMO assessment and support prioritizing strict control of light exposure over many other real-world contextual variables when the primary objective is characterization of the endogenous circadian phase [10, 15–17, 22, 36].

The present study also demonstrated substantial adaptability of circadian timing following sustained behavioral delay of the sleep-wake schedule. After a 10-day, 5-hour delay in habitual bedtime, DLMO timing shifted later in approximate alignment with the delayed behavioral schedule while preserving overall melatonin profile morphology. Following resumption of the participant’s habitual sleep schedule, circadian phase returned toward baseline over several days, suggesting preserved responsiveness to behavioral (primary) and environmental (secondary) entrainment cues. These observations illustrate the dynamic interaction between behavioral sleep scheduling and endogenous circadian physiology in an individual exhibiting otherwise remarkable long-term circadian stability [26].

These findings raise an important translational question. Although delayed circadian phase is often viewed as a relatively fixed characteristic, this case demonstrates that substantial phase delays can arise through behavioral manipulation alone in an individual with an otherwise highly entrained circadian system. This suggests that similar circadian phase presentations may reflect fundamentally different underlying mechanisms, with some individuals exhibiting preserved entrainment capacity and others demonstrating impaired responsiveness to environmental or behavioral zeitgebers. Future studies examining individual differences in circadian adaptability and re-entrainment may help refine the biological characterization of delayed circadian endotypes beyond phase timing alone [24, 25].

## CONCLUSIONS

At-home salivary DLMO assessments demonstrated substantial intra-individual stability across baseline conditions, supporting the feasibility of home-based circadian phase characterization when standardized protocols are followed. Evaluated against a robust longitudinal baseline, several commonly cited contextual factors produced relatively small deviations from baseline, whereas bright-light exposure and recent melatonin supplementation substantially altered the interpretability of DLMO assessments. These findings illustrate how contextual influences reported in the literature may present within a well-characterized individual and provide preliminary insight into their relative magnitude compared with normal intra-individual variability. Although confirmation in larger cohorts is needed, this work represents an initial step toward distinguishing contextual factors that warrant stringent protocol control from those that may contribute only modest variation, with the potential to reduce patient burden while preserving the translational and clinical utility of at-home DLMO assessment.

## DECLARATIONS

Ethics approval and consent to participate: The participant provided informed consent for inclusion and publication of de-identified results.

Funding: No external funding.

Competing interests: Authors affiliated with Salimetrics LLC have potential commercial interests related to circadian phase assessment services.

Data availability: De-identified data supporting the findings are available from the corresponding author upon reasonable request.

## Data Availability

All data produced in the present work are contained in the manuscript

